# A Prototype Machine Learning Pipeline for Assessing and Tracking Keloid Scars

**DOI:** 10.1101/2024.09.30.24314501

**Authors:** Mahla Abdolahnejad, Armita Zandi, Jordan Wong, Hannah O. Chan, Victoria Lin, Hyerin Jeong, Rakesh Joshi, Joshua N. Wong, Colin Hong

## Abstract

Dysregulated wound healing, marked by excessive collagen deposition, is the hallmark to keloid scar formation. Current methods for assessing keloids in clinical settings rely heavily on subjective measures, which are prone to interrater variability. This study introduces a machine learning (ML) pipeline prototype, designed to automate the detection, measurement, and colour analysis of keloid scars. Using a convolutional neural network (CNN), the pipeline segments keloid lesions from 2D images, applies fiducial markers for accurate size measurement, and utilizes K-Means clustering for colorimetry analysis. The CNN achieved a classification accuracy of 98% on a small test dataset. Segmentation was further refined using binary masks and contour-based detection.

Colorimetry analysis revealed heterogeneity in pigmentation across keloid lesions, was varied by patient skin type, and tracked changes over time. The pipeline was validated on patients over a 5–6-month period, accurately detecting changes in keloid size and colour. While the algorithm was highly effective in most cases, challenges were noted in patients with nascent keloid or those with dark skin tones where the contrast between keloid and skin was insufficient for accurate segmentation. Additionally, early-stage keloid detection showed inconsistencies in defining lesion boundaries, particularly when keloids expanded rapidly. Despite these limitations, the ML pipeline presents a promising tool for objective keloid assessment, offering a practical, accessible, and accurate alternative to current clinical practices.

## Introduction

Skin is the largest organ in the human body, acting as the first physical barrier providing protection from infection, dehydration, and mechanical forces. Whilst providing protection, skin has a paramount role in sensation and mobility with dynamic elastic properties that enable a full Range-of-Motion (ROM) through the body^1^. Damage to the skin can disrupt many of these functions and lead to variable outcomes based on injury- and host-specific factors. The depth, mechanism, and location of the injury contributes to scar formation at the key phases of wound healing: hemostasis, inflammation, proliferation, and remodeling^2^. Goals following injury involve returning baseline function, ameliorating associated pain, and improving aesthetic outcomes. Surveys assessing the impact of scars have shown significant reduction in quality-of-life related to limitations in function (reduced ROM), sensation (pain and pruritus), and appearance, with the latter being the most frequent concern^3,4^. The effects of visible scars impairs psychosocial well-being, spanning subjects’ professional to social and intimate lives, and puts them at a higher risk of social exclusion^5^. As such, there are major strides to improve outcomes with management strategies that vary with scar type.

Keloid scars, consisting of a disordered healing matrix, is hypothesized to involve increased collagen and extracellular matrix synthesis and deposition. Following trauma to the skin, keloidal fibroblast cells proliferate and persist excessively when compared to non-pathological wound healing. This results in elevated collagen and proinflammatory cytokine production^6^. While the exact pathophysiology remains unknown, the imbalance between the ratio of collagen deposition to degradation is characteristic of keloids with a continuation of deposition beyond the typical wound repair period ^7,8^. This dysregulation in collagen synthesis leads to the clinical presentation of a raised lesion that extends beyond the initial injury site. Invasion into adjacent healthy skin along with tension due to reduced keloid elasticity may impair a patient’s movement. Furthermore, the pattern of keloid growth along the plane of tension (e.g., horizontal growth of a lesion across the anterior chest) further limits patient function^9^. Keloid therapies aim to resolve scar contractures, reduce size/thickness and progression, and improve discolouration to help return baseline function. Management of pruritus and pain reported with keloids is an additional therapeutic goal^10^. In brief, therapies employed include corticosteroid injections, laser ablation, surgical excision, cryotherapy, and radiotherapy^6-8^. While many of these management strategies require multiple treatments, determining if a lesion is responsive to a specific treatment method is imperative to prevent unnecessary treatment, reduce costs, and hasten patient recovery.

Across Canada, there is no standardized approach to assess scars in clinical practice. Current gold standard measurements (e.g., modified Vancouver Scar Scale [mVSS]^11^, Patient and Observer Scar [POSAS]^12^) utilize scoring systems that are based on subjective interpretation of visual and tactile properties. Issues within these subjective scales arise with interrater variability and patient bias. Alternatively, objective measurement techniques such as ultrasound, cutometry, and mexametry can report quantitative data, such as vascularity, colour, and elasticity. However, quantitative analysis require validation with qualitative scales^11^, and the tools required to obtain quantitative measures are cost-prohibitive and impractical in clinical encounters. Patients with geographic or financial barriers face additional hurdles which prevent adherence to regular follow-up appointments. As such, the gap in clinical practices necessitates a scar assessment tool that produces objective measures and is easily accessible, affordable, and accurate.

Machine learning (ML), particularly computer vision systems that use convolutional neural networks (CNN), with architectures like EfficientNet B7, have been shown to be well adapted for classification, segmentation, and visual attribute assessment tasks for skin-related conditions ^13,14,15^. These systems have the potential to enhance the identification of keloids from other skin lesions by leveraging its advanced neural network architecture for feature extraction^13^. EfficientNet B7 utilizes a compound scaling method that balances depth, width, and resolution, providing superior accuracy and efficiency in image classification tasks^16^. Thus, this model can be trained on small keloid image datasets, allowing it to learn and identify the distinct visual attributes of keloidal scars, such as their texture, colour, and shape. Compared to subjective human analysis, EfficientNet B7 reduces variability and potential errors introduced by human observers, leading to more consistent and accurate identification.

Incorporating activation channel maps and saliency maps, products of a CNN’s “seeing” and “understanding” processes, can further refine the segmentation and contouring of keloids. Activation channels help identify which parts of the network are most active when processing an image, providing insights into features that are most relevant for distinguishing keloids. Saliency maps highlight areas of an image that most influence the model’s predictions, which enables precise localization of keloidal regions^14^. These ML tools generate detailed segmentation maps, delineating the boundaries of keloids accurately. This process is crucial for effective treatment planning and monitoring the progression or regression of keloids over time.

OpenCV, a powerful open-source computer vision library, can be used to measure the size of keloids using edge detection and fiducial markers for size reference. By placing a known-size marker in the image, OpenCV can calibrate the measurements, ensuring accurate size determination of keloidal scars. Additionally, algorithms like K-Means clustering can be used to analyze and quantify the colour distribution within a keloid segment. This algorithm groups pixels based on their colour similarity, providing a detailed colour profile of the keloid, which is useful for assessing changes in pigmentation and vascularity over time^12,15,17^. These techniques utilized in a single machine learning pipeline form a comprehensive approach to automate the analysis and monitoring of visual aspects of keloids, and enhance clinical outcomes and objective research capabilities. In this proof-of concept study, we describe the methodology and preliminary results, using actual keloid patient images, allowing time-series tracking of size, shape and colorimetry.

## METHODS

### Machine Learning

#### Machine Learning pipeline

The keloid machine learning pipeline consists of several key components (Figure 1): image preprocessing, feature extraction, classification, segmentation, contour/boundary detection, and colorimetric analysis. The first stage involves a CNN classifier, trained to distinguish between 2D color images of three classes: keloids, skin lesions, and other dermatological conditions. The activations from various layers of the trained classifier are used to generate a visualization map that highlights areas affected by keloid lesions. Once this map is created by extracting features from the CNN, segmentation of the keloid lesions is performed using a thresholding approach based on pixel intensity. This method enables the precise identification of keloids and delineation of their contours. The next component utilizes image processing libraries such as OpenCV (Open-Source Computer Vision Library) to measure the size of the detected keloid lesions using a fiducial marker of known dimensions. This marker is detected through contour-based methods, allowing for an accurate pixel-to-millimeter conversion. Additionally, the fiducial marker can be used to calibrate the system for colorimetric analysis. Finally, a K-Means clustering algorithm is applied to analyze the color of the lesions within the segmented areas, determining the optimal number of color clusters for pigment analysis. This comprehensive pipeline processes and evaluates keloid characteristics across multiple dimensions.

**Figure 1:**
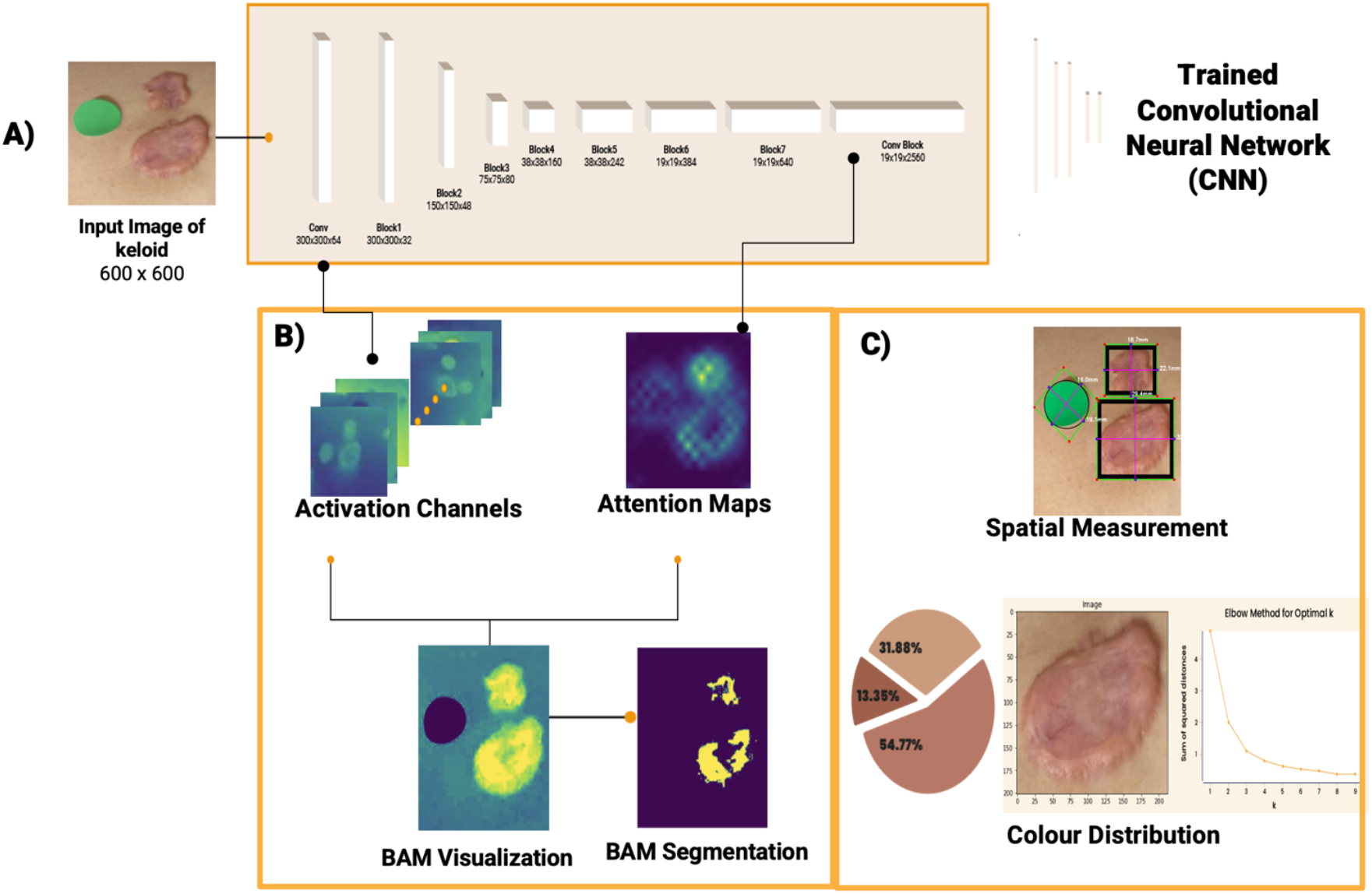
Deep learning-based analysis of skin lesions using a Convolutional Neural Network (CNN) **A)** Input image of a skin lesion (600×600 pixels) is processed through a trained CNN architecture. **B)** Visualization of the CNN’s internal representations: Activation channels highlight relevant features detected by the network; Attention maps show areas of focus for the model’s decision-making; BAM (Boundary Attention Map) Visualization displays a heatmap of the most salient regions; BAM Segmentation outlines the predicted lesion boundaries. **C)** Quantitative analysis of the lesion: Spatial Measurement conducted using bounding box indicating the lesion’s location and size; Keloid colour distribution is depicted Pie chart showing the proportion of different colour components within the lesion. Colour cluster identification elbow graph

#### Training of a Convolutional Neural Network

A CNN model based on a pre-trained EfficientNetB7 was utilized, with modifications to the classifier head to include additional fully connected layers and a 3-class SoftMax layer for keloid classification from 2D images. The modified CNN was fine-tuned on 6,550 2D color images (including augmented images) of keloid patients, using 5-fold cross-validation and categorical cross-entropy as the loss function. The final architecture was obtained through hyperparameter optimization, which included adding 4 fully connected layers, adjusting learning rates, and optimizing regularization via dropout variables, among other parameters.

The model’s performance during training was evaluated based on increases in categorical validation accuracy and/or decreases in validation loss. Hyperparameter optimization was iteratively refined over multiple training rounds. A detailed statistical analysis of the model’s performance metrics—accuracy, loss, precision, recall, and F1 score (harmonized accuracy) was conducted using an additional set of 219 images as a validation set. A separate test set of 63 images was employed to assess the model’s final performance. Backgrounds in these images were removed by masking non-skin objects within the image field.

#### Segmentation of Keloids

Keloid segmentation was performed using a binary mask-based approach. After generating a visualization or heatmap that highlights the keloid areas, it is used to create segmentation masks. Initially, a Gaussian Mixture Model (GMM) is applied to the pixel values of the heatmap, and the intersection points of the Gaussian components are calculated to segment the heatmap into a binary mask. The resulting binary segmentation mask then undergoes post-processing to filter out noise. Smaller artifacts are removed, and the edges are smoothed to improve accuracy using Boundary Attention Maps (BAM)^14^. This method refines the segmentation by applying contour extraction and filtering based on a custom criterion to enhance the precision of the output.

Finally, a contour detection technique is applied to extract the keloid lesion’s boundaries. Bounding boxes are drawn around the segmented keloid areas, ensuring accurate demarcation. This method was consistently applied across multiple patient images, ensuring reliable detection of keloid boundaries across different lesion sizes and varying skin tones.

#### Measurement of Keloids using a Fiducial Marker

As a reference for measurement, a round fiducial marker with a diameter of 19.1 mm was placed near the keloid. This fiducial marker, with its known size, enabled the conversion of pixel measurements to real-world dimensions. After segmenting the keloid using a binary mask, contours were drawn around the segmented areas. For each contour, a minimum-area bounding box was calculated, and its width and height were measured in pixels. These pixel measurements were then converted to actual dimensions using the fiducial marker as a reference. The bounding boxes were overlaid on the images to visually confirm the keloid’s dimensions.

#### Using K-Means Clustering Algorithm for Keloid Colorimetry

The K-Means clustering algorithm was applied to analyze the colour distribution of keloid lesions. The input keloid images were first cropped to isolate the keloid area, and the segmented regions were processed by reshaping the image data into an array of RGB values. To exclude non-lesion pixels, a binary mask was applied as described in the previous sections. After preprocessing, K-Means clustering was used to group the lesion pixels based on their colour values. The elbow method was employed to determine the optimal number of clusters by plotting the sum of squared distances for clusters ranging from 1 to 10. The optimal number of colour clusters was identified programmatically. The K-Means algorithm then assigned each pixel to one of the detected colour clusters, and the proportion of pixels in each cluster was computed. The results were visualized with a pie chart showing the percentage of pixels corresponding to each colour. This colorimetry analysis provides insights into pigmentation changes within the keloid, tracked over time

### Proof-of-Concept: Tracking patients

As proof-of-concept, the pipeline was tested by tracking the progression of the keloid scars on patients (Patient1 and Patient2) over the course of 5 and 6 months, respectively. High resolution images of the keloids with a fiducial marker were captured with a smartphone camera (iPhone 8, iOS device), and were ingested by the algorithm to detect changes in scar measurement, ratio of colours, and contour of the keloids over a span of 5 timepoints.

## RESULTS

### CNN-based classifier Performance

The pre-trained CNN-based keloid classifier, using and EfficientNet B7 architecture, was trained on a dataset of 6550 2D colour images, with 219 images used for validation and 63 images for testing. The frozen model accuracy reached approximately 97.5% for the training set and 95% for the validation set (Figure 2A). In comparison, the standard, unfrozen model achieved near-perfect accuracy, reaching 100% on the training set and 98% on the validation set (Figure 2B). The classifier demonstrated robust performance in distinguishing between keloid, normal, and other skin images, achieving test set accuracies of 100%, 97%, and 100%, respectively. The recall scores for keloid, normal, and other images were 94%, 100%, and 100%, respectively, as shown in the confusion matrix (Figure 2D). The overall F1 scores for keloid, normal, and other images were 97%, 98%, and 100%, with an overall F1 accuracy of 98%. The macro averages for precision, recall, and F1-score were 99%, 98%, and 98%, respectively, with weighted averages of 98% across all metrics.

**Figure 2:**
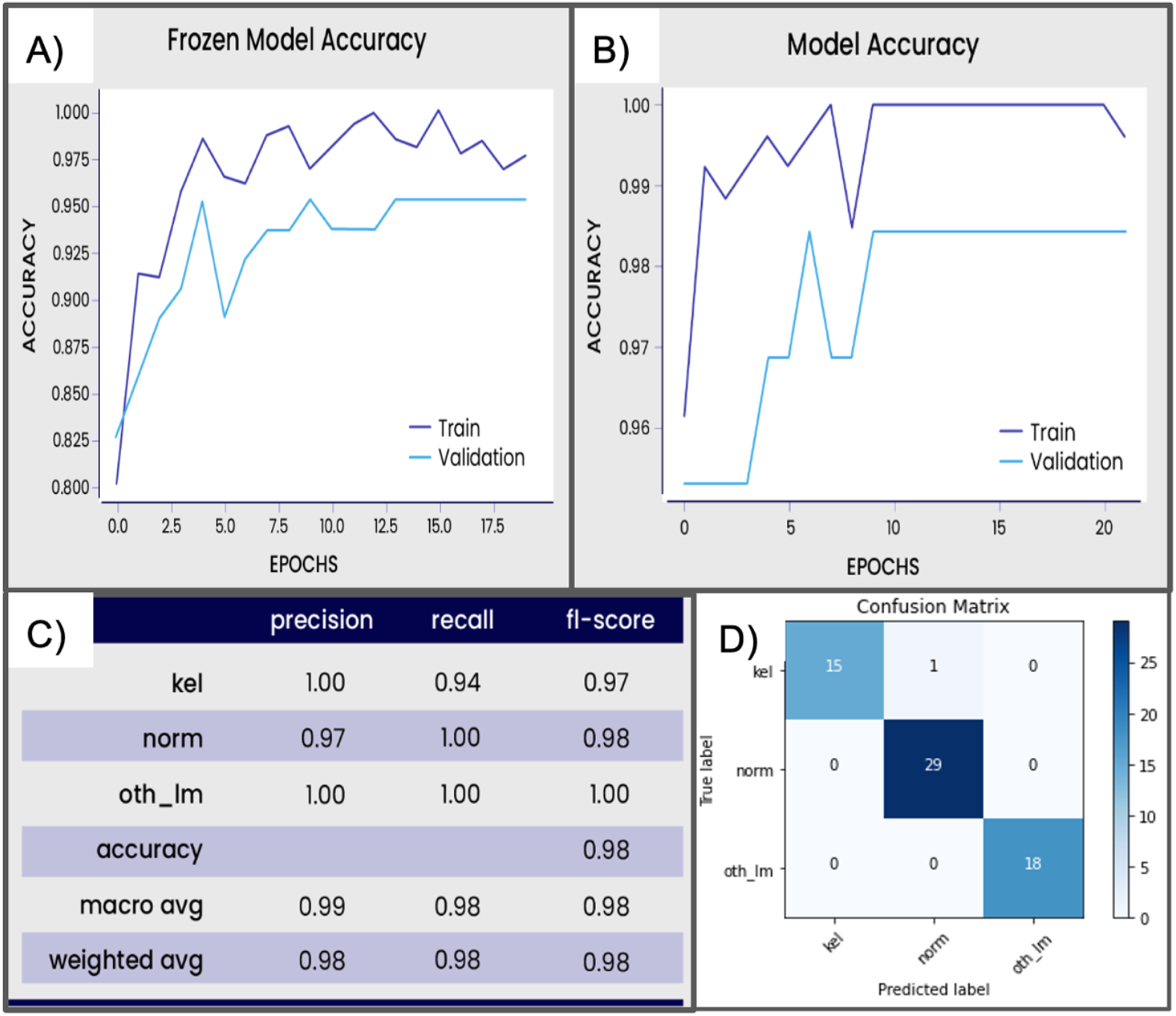
Performance analysis of a Keloid classification model using EfficientNet B7 architecture. **A, B)** Frozen Model and Final Model graphs showing the training and validation accuracy over training epochs for the model with frozen (A) and unfrozen (B) layers of the B7 architecture. The blue line represents training accuracy, while the light blue line shows validation accuracy. **C)** Performance Metrics Table enumerates Precision, recall, and F1-score for different classes (keloid, normal skin, other lesions), Overall accuracy and Macro and weighted averages of the metrics. **D)** Confusion Matrix that visualizes the model’s predictions versus actual labels. The diagonal elements represent correct classifications, while off-diagonal elements show misclassifications.

### Measurement and Segmentation Analyses

To test the quality of the segmentation and size measurement, images with fiducial marker present were used for the keloid measurements of the initial input image, as well as a preliminary segmentation of the keloid scars, as shown in Figure 3. Figure 3 A, D and G are the input images from three different Keloid patients (Patient1, Patient2 and Patient3, respectively) who have various sized keloids, skin tones, and scar colourations, that were captured during their first clinic visit. The segmentation and measurement components of the AI pipeline segmented the scars with a high precision, although there were confounding factors when measuring larger complex keloid scars. For example, the four discrete keloids were measured from the first patient (Figure 3B), with dimensions of Keloid 1: 37.7 mm x 4.2 mm; Keloid 2: 10.2 mm x 5.1 mm; Keloid 3: 8.3 mm x 6.5 mm, and Keloid 4: 10.4 mm x 3.8 mm. The segmentation was carried out with fine granularity in depicting the boundaries of the keloid scars (Figure 3B, C). On the other hand, the algorithm measured the prominent features of the second patient’s complex single scar (Figure 3D) as multiple discrete scars (Figure 3E), even though the boundary segmentations were accurate (Figure 3 E, F). It was also observed that indented scars with varied colouration (Figure 3G) can be sized (Figure 3H) and segmented accurately (Figure 3I). This method was applied to several keloid images, yielding accurate measurements of lesion size with a high precision of ± 2 mm, despite several complex scars being identified as discrete scars and again measured with high precision for each individual scar. One observed limitation in the segmentation algorithm was inaccurate measures for our patient with a dark Fitzpatrick Type VI skin tone who had and keloid scars with a similar skin tone (Supplemental Figure 1).

**Figure 3:**
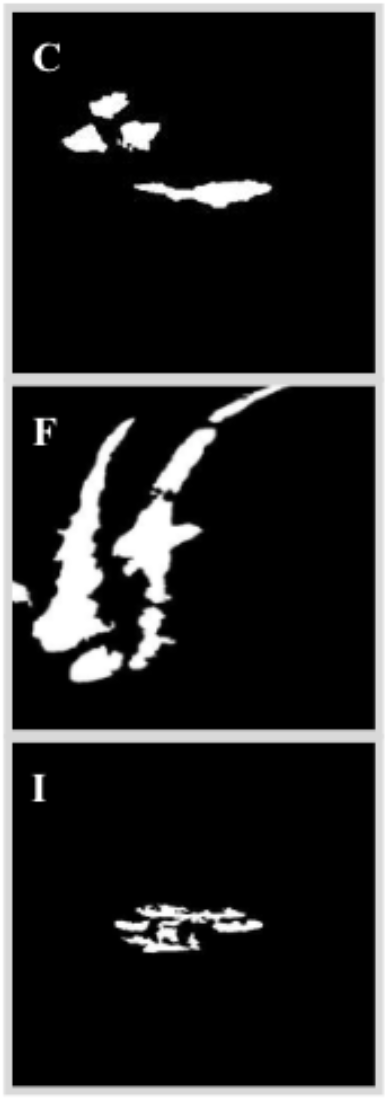
**(A, D, G)** Input images of patients presenting with keloid scars on various body regions. A fiducial marker (green or yellow circle) is placed adjacent to the scar to provide a reference for scale and accurate measurement. **(B, E, H)** Scar dimensions are delineated using an automated segmentation algorithm that utilizes the fiducial marker for calibration. The segmented regions of interest (ROI) corresponding to the keloid scars are outlined in blue. **(C, F, I)** Binary masks generated from the segmentation process, where the scarred areas are represented as white regions. These masks are used for further quantitative analysis, including evaluating the scar’s size and shape

### Colorimetric Analyses

Segmentation and colorimetry were performed on the same keloid images from the three patients (Figures 4 A, D, G). The keloid regions were accurately segmented from the surrounding skin using binary masks (Figure B, E, H). Post-segmentation, K-Means clustering was applied to the keloid pixels to identify and categorize the dominant colours within the scars.

**Figure 4:**
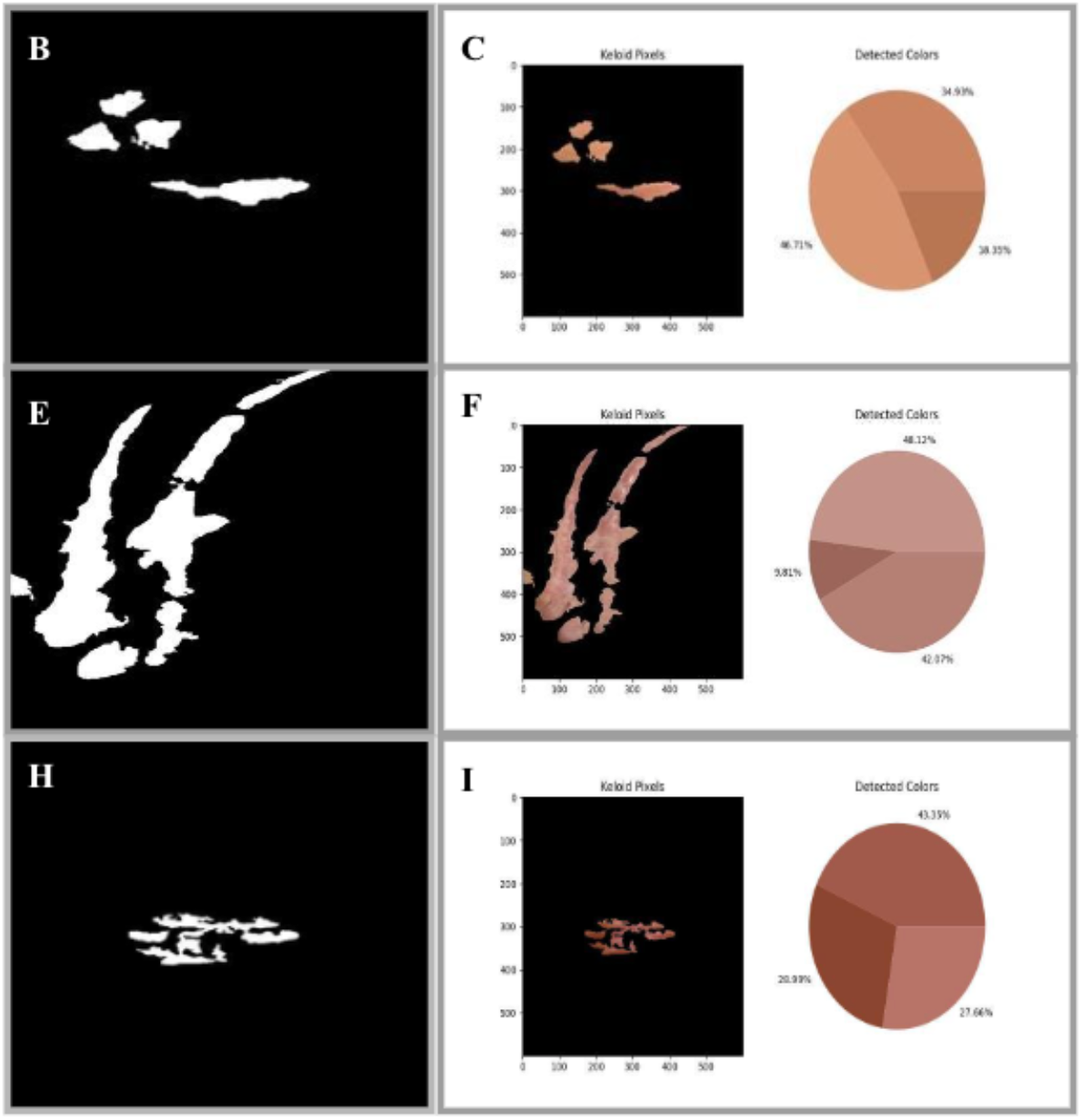
**(A, D, G)** Input images of keloid scars from three patients with fiducial markers for reference. **(B, E, H**) Binary masks accurately segment keloid regions from surrounding skin for further analysis. **(C, F, I)** K-Means clustering applied to the segmented keloid pixels identifies three dominant colour clusters, shown in pie charts. For Patient 1 **(A-C)**, 46.71% of the keloid was lighter pigmentation; for Patient 2 **(D-F)**, 48.12% was light reddish; and for Patient 3 **(G-I**), 28.77% was the darkest tone.

For Patient1(Figure 4A-C), the colorimetry analysis identified three main colour clusters, with 46.71% of the lesion being classified as a lighter pigmentation and 18.35% as the darkest shade (Figure 4C). Classified as skin type III on the Fitzpatrick scale, this patient’s keloids were clinically determined to be a beige-brown tone and demonstrated higher erythema and lower melanin levels.

In Patient2(Figures 4 D-F), the analysis revealed three distinct colour clusters, with the majority (48.12%) of the pixels corresponding to a light reddish colour, 42.07 % being darker, and 9.81% representing the darkest shade (Figure 4F). This patient, with a Fitzpatrick skin type II, had darker keloid pigmentation due to higher melanin levels in the scar compared to their natural skin tone.

Patient3 (Figures 4 G-I), classified as Fitzpatrick skin type IV, exhibited three primary colour clusters, with 27.66% of the lesion represented by a lighter tone and 28.77 % by the darkest one (Figure 4 I). The detected colours for this patient’s keloids were darker shades, contrasting with the patient’s skin tone and reflecting higher melanin content.

### Tracking a Keloid over time

Figure 5 shows the results of tracking a single patient’s keloid cluster over time. The patient had distinct keloids on the left side of her face which visibly darkened and increased in size over the course of 5 months (Figure 5 Panel A). The algorithm supported the visual progression of the size and darkening of the colour over time and accurately identified the keloid area each time at 5 time points (Figure 5 Panels B, C, Table1). Interestingly, the algorithm failed to recognize the entire keloid located on the bottom of the patient’s face. However, the algorithms successfully defined the borders of the same keloid t in the following timepoint, indicating that this error probably occurs on recently expanded keloids (Figure 5 Panel B, Panel III). Additionally, the algorithm failed to define the keloid area on darker skin tones presumably due to the lack of contrast in pigment between the patient’s skin tone and the keloid (Supplemental Figure 1).

**Figure 5:**
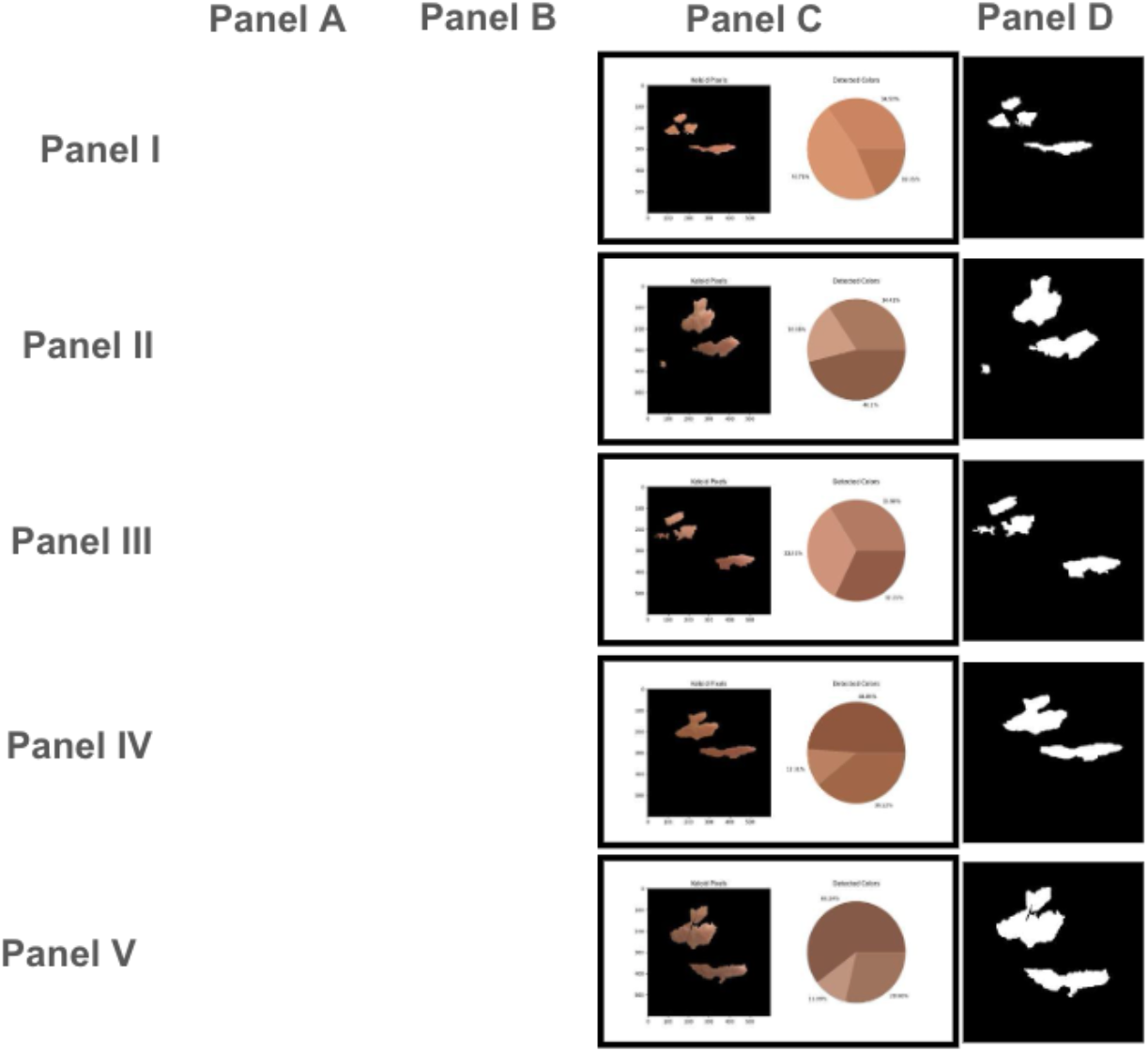
Algorithms were used to track a single patient with keloid scar cluster on cheeks(Patient1). Composite figure shows the progression of a single patient’s keloid clusters over five months. Panel A displays input images of the keloids on the left side of the patient’s face, which visibly darkened and expanded over time. Panel B shows the algorithm’s segmented keloid regions, while Panel C highlights colour analysis via pie charts at each time point. Panel D presents the binary masks used for area detection. The algorithm successfully tracked keloid size and darkening but initially missed part of the bottom keloid, which was correctly identified in later time points (Panel B, III). Dark skin tone challenges in keloid detection.

Table 1 summarizes the results of tracking a single keloid on two patients over a 5- and 6-month period. Changes in size (in mm) and colours detected by the algorithms described indicate no change in the colour composition in the keloids of both these patients but detected a change in the size of the keloids. Tracking of colour changes was made possible through the tracking of value attributes for each colour (Ex: hexadecimal or RGB values). Patient1’s a cheek keloid (Fig 5 Panel1) doubled in size over a 4-month period, from 37.7mm x 4.2mm (158.34 mm^2^) to 21.0 mm x 16.8 mm (366.24 mm^2^). Patient2’s upper back keloid (see Figure 4G) increased rapidly from 11.6mm x 2.1mm (24.36 mm^2^) to 19.3mm x 8.7 mm (167.91 mm^2^) over a 6-month duration, effectively increasing over 6 times from its initial size. A limitation of the size algorithm longitudinal tracking of keloids exists in smaller keloids. Through extensive testing of the size algorithm, an error rate of ±2 mm was found. Therefore, this results in higher error rates for smaller keloids that are measured and segmented, especially early in its presentation

**TABLE 1:** Tracking size changes and colours detected in a single keloid, in 2 patients Patient1 and Patient2, over 5 timepoints (5 and 6 months, respectively). Timepoints, when image data was not collected or there was an error in the algorithm correctly identifying the entire tracked keloid, were excluded from the analysis (N/A value).

|  | Patient 1 |  | Patient 2 |  |
| --- | --- | --- | --- | --- |
|  | Keloid Size( in mm) | Keloid colours | Keloid Size(in mm) | Keloid colours |
| TIMEPOINT 1 | 37.7 x 4.2 | 3 | 11.6 x 2.1 | 3 |
| TIMEPOINT 2 | 31. x 11.7 | 3 | N/A | N/A |
| TIME POINT 3 | N/A | N/A | 14.9 x 3.3 | 3 |
| TIMEPOINT 4 | 24.0 x 9.2 | 3 | N/A | N/A |
| TIMEPOINT 5 | 21.0 x 16.8 | 3 | 19.3 x 8.7 | 3 |

## Discussion

Tracking a keloid’s visual and tactile properties over time is the primary way clinicians discern lesion growth to regression. In addition to patient feedback, examining the size, pigmentation, vascularity, pliability, and scar height can dictate the type of treatments received or discontinued^11,12^. Studies that compare a mainstay treatment of steroid injections to alternative therapies (CO2 laser therapy^18, 19^, cryotherapy, brachytherapy^20^) primarily used the subjective scales (POSAS and VSS) to determine treatment efficacy.

Although clinically practical, use of these assessments leaves room for interrater variability and bias which prevents robust conclusions to be drawn from clinical trials. Those that also use objective measures, such as keloid volume determined with plaster molds^20^ are labour intensive. These drawbacks, reflected in clinical practice, necessitates the creation of a practical and accurate objective assessment tool. In the present study, we aimed to address this with our novel CNN used to examine keloids via smartphone images.

In the present proof-of-concept study, our novel ML pipeline and fiducial marker approach showed promise in three domains: 1) identification of keloid scars with high accuracy, precision, and recall; 2) detection of differences in colour pigmentation and formulating an output of keloid colour distribution; and 3) measurement of the lesion size with an error rate of ±2 mm, clinically correlated with manual measurements. Two shortcomings of this algorithmic pipeline were also seen such as higher error rates for small keloids and the lack of colorimetric values to understand the change in colour composition. Heterogeneity in pigmentation within keloid lesions was discovered which highlights the potential of colorimetric analysis to quantify subtle variations in skin pigmentation across patients. This also highlights a short coming of the study as cross-referencing values for erythema and melanin for different shades was not performed which is measured by medical devices such as mexameters. This can be done using hexadecimal or RGB values for erythema and melanin counts that translate to the validated erythema and melanin indices used by clinicians.

We demonstrated the utility of our algorithm in tracking developing keloids over 4–6-month periods. In the early stages of keloid development, the algorithms failed to capture the entire lesion leading to inaccurate size measurements and boundary identification. This may be attributed to the distortions in the image or the expansion of the keloid. The AI may have failed to capture the recently expanded area of a keloid when the keloidal area is not pigmented or raised like mature keloids. This was confirmed with correct contour mapping of the same keloid in the following timepoints, indicating that this error only occurs on very recently expanded keloids. Additional tracking of immature keloids is needed to confirm this hypothesis and improve detection with machine learning. When applied to more mature keloids, the entire lesion was detected by the algorithm and the outputs in size and colour were easily compared. This data can help indicate keloid progression or regression and direct patient care.

We believe that this proposed this ML algorithm, with modifications to overcome shortcomings, as an objective measurement tool to complement clinician observation. There are ongoing efforts to create objective keloid assessments include molding^21^, sensing devices^22^, and three-dimensional imaging (stereophotogrammetry)^23^. Despite their promise, these approaches are not widely accessible across centers and may not be practical in every clinical encounter. Contrarily, the present study’s d tool which is adapted for smart devices not only increases accessibility across treatment centers but increases accessibility for patients who face barriers in attending in-person appointments. Smart device pictures are regularly taken for patient documentation, thus integrating our algorithm into a clinical workflow would prove practical. Furthermore, our technology may be utilized in a virtual care model if the patient has access to a smart device and an inexpensive fiducial marker. With all considered, following the progression of a keloid may become more feasible with concrete objective readings by ML algorithms.

Limitations in the present study relate to validation and the nature of the approach. While the focus was to provide a proof-of-concept for AI in scar assessment, we did not compare outputs of the algorithm to other objective tools, and on a larger population. Sensing devices that examine pigmentation could also be used to compare with our ML colour analysis outputs. The Mexameter® is a reliable and precise differentiator of the colour properties (erythema and pigmentation) in burn scars^24^. Comparing the algorithm to readings of the Mexameter® can help validate the colour analysis feature of our approach. Because only 2D images were used to train the algorithm, examining scar volume and height would be challenging. Building on our concept, we aim to expand scar analysis with video captures which may overcome the limitations of 2D analysis

Overall, we propose that ML pipelines can be a useful objective tool to serve alongside clinician observation in keloid assessment. These measurements can overcome issues of bias and interrater variability when tracking the progression of a lesion. More work needs to be done in further developing the algorithm to improve accuracy and to broaden the type of measurements that can be generated, such as its application to other scar types, such as hypertrophic scars due to burn injuries. Our work supports further efforts to integrate ML and Mobile Technology in clinical scar assessment.

## Data Availability

All data produced in the present work are available upon reasonable request to the authors

---

**Supplemental Figure 1:** Limitations of the segmentation algorithm in recognizing keloid scars in darker skin toned patients

